# Length of ischemic time is critical for accurate determination of homologous recombination capacity by immunostaining in FFPE tumor samples

**DOI:** 10.1101/2024.11.28.24318148

**Authors:** Ezgi Karagöz, Sanna Pikkusaari, Manuela Tumiati, Anni Virtanen, Matilda Salko, Anni Härkönen, Anna Kanerva, Heidi Koskela, Johanna Tapper, Riitta Koivisto-Korander, Titta Joutsiniemi, Ulla-Maija Haltia, Heini Lassus, Anniina Färkkilä, Johanna Hynninen, Sakari Hietanen, Liisa Kauppi

## Abstract

Homologous recombination-deficient (HRD) high-grade serous ovarian cancers (HGSC) are more sensitive to PARP inhibitors compared to their homologous recombination-proficient counterparts. To match the right drug with the right patient the HRD status must be accurately determined. Functional HRD assays, which assess HRD status by quantifying RAD51, a key homologous recombination (HR) protein, are a promising approach for identifying HRD cases. However, these tests are yet to be optimized for pre-analytical variables, specifically HGSC tissue sampling protocols, which can impact RAD51 signal measurement. In this study, we systematically analyzed the impact of ischemic time on formalin-fixed paraffin-embedded HGSC specimens. We demonstrate that the maximum length of ischemic time compatible with accurate HRD calls is 2 hours post-excision. Our findings highlight the importance of properly monitoring and recording sample handling processes, particularly in HGSC, and warrant caution when using archival tumor material where this information is unavailable. Non-optimal pre-analytical factors like ischemic time can cause false HRD calls, thus leading to incorrect patient stratification, which may result in the initiation of treatments with potential side effects without a therapeutic benefit.

## Introduction

In the past five years, treatment for high-grade serous ovarian cancer (HGSC) has been transformed by the introduction of Poly ADP-ribose polymerase (PARP) inhibitors as maintenance therapy, with homologous recombination deficiency (HRD) emerging as a key biomarker for predicting PARP inhibitor sensitivity and chemosensitivity (Coleman 2017). HRD cases cannot overcome double-strand breaks, making them targetable by DNA damaging agents such as platinum and PARP inhibitors. In contrast, approximately 50% of HGSC cases have homologous recombination proficient (HRP) tumors, which are linked to primary resistance to platinum-based chemotherapy and PARP inhibitors as the cells can repair the DNA damage, resulting in poorer survival outcomes. The classification of HRD cases in the clinic currently relies on genomics-based approaches. However, The European Society for Medical Oncology (ESMO) has advocated for the utilization of biomarkers to predict treatment outcomes and determine HRD status, aiming to assess the patients likely benefit from PARP inhibitors (Miller 2020).

Existing HRD assays fall into three main categories: (1) analysis of homologous recombination repair (HRR) gene mutations such as BRCA1/2, (2) genomic “scars” and mutational signatures, and (3) functional HRD tests (Chiang 2021, Pennington 2014). Successful DNA-based HRD testing typically requires at least 30% tumor cells in the sample and sufficient total tumor material for DNA extraction; these conditions are not always met, especially for tumors of patients treated with neoadjuvant chemotherapy or for needle biopsy samples. By contrast, functional HRD testing can be performed even with scarce tumor material. It assesses a tumor’s homologous recombination (HR) status by quantifying RAD51, a central HR protein, in tumor nuclei (Pikkusaari 2023, Castroviejo-Bermejo 2018, Blanc-Durand 2023, van Wijk 2021, Compadre 2023) from formalin-fixed paraffin-embedded (FFPE) specimens. A significant advantage of functional HRD testing is that it can be performed on formalin-fixed paraffin-embedded (FFPE) specimens that are collected as part of routine clinical diagnostics, even though the tumor material in FFPE block is limited.

We previously demonstrated that the functional HR (fHR) assay, which measures HR capacity by quantifying RAD51 foci in S-phase cells, predicts sensitivity to platinum-based chemotherapy and PARP inhibitors in HGSC (Pikkusaari 2023). However, in the context of the same study we noted that tissue handling processes, particularly from surgical excision to sample fixation, appeared to be critical for yielding reliable fHR results. Specifically, we observed that samples with prolonged ischemic times (time from resection to fixation of a tissue specimen) had less RAD51-positive nuclei (Pikkusaari 2023). Loss of protein signal for markers like HER2, ER, PR, and Ki-67 upon extended time is well-documented in breast cancer (Hammond 2010, Neumeister 2012, Yildiz-Aktas 2012, Khoury 2018). Accordingly, the American Society of Clinical Oncology/College of American Pathologists (ASCO-CAP) guidelines state that breast cancer tissue should be fixed within one hour to prevent ischemia, acidosis, and enzymatic degradation (Hammond 2010). In contrast, sampling protocols for HGSC are currently not standardized or optimized for ischemic time. Functional HRD testing has a future niche in clinical diagnostics, predicting PARP inhibitor response, and therefore it is important to delineate the effects of delayed time-to-fixation, particularly on nuclear RAD51 signal. In this study, we systematically examine the impact of ischemic time on the HR biomarker RAD51 protein, in HGSC samples.

## Materials and Methods

### Patients and tumor specimens

All HGSC specimens in this study were obtained from participants who gave written informed consent. The study was approved by the ethics boards of the Wellbeing Services county of Southwest Finland (DECIDER study, clinical trial identifier NCT04846933, ethics board approval VARHA/28314/13.02.02/2023) and the Helsinki University Hospital (HUH) ONCOSYS-OVA study, clinical trial identifier NCT06117384, and the use of samples and clinical data was approved by the local Ethics Committee, HUS334/2021).

All experiments followed the ethical guidelines of the WMA Declaration of Helsinki. Tumor specimens used in this study were resected from the following anatomical sites: ovary, omentum, fallopian tube, and peritoneum. This study utilized a total of 27 pre-treatment specimens from 24 HGSC patients forming two cohorts, referred to as retrospective and prospective, to examine the impact of ischemic time on fHR testing (Figure 1A).

**Figure 1.**
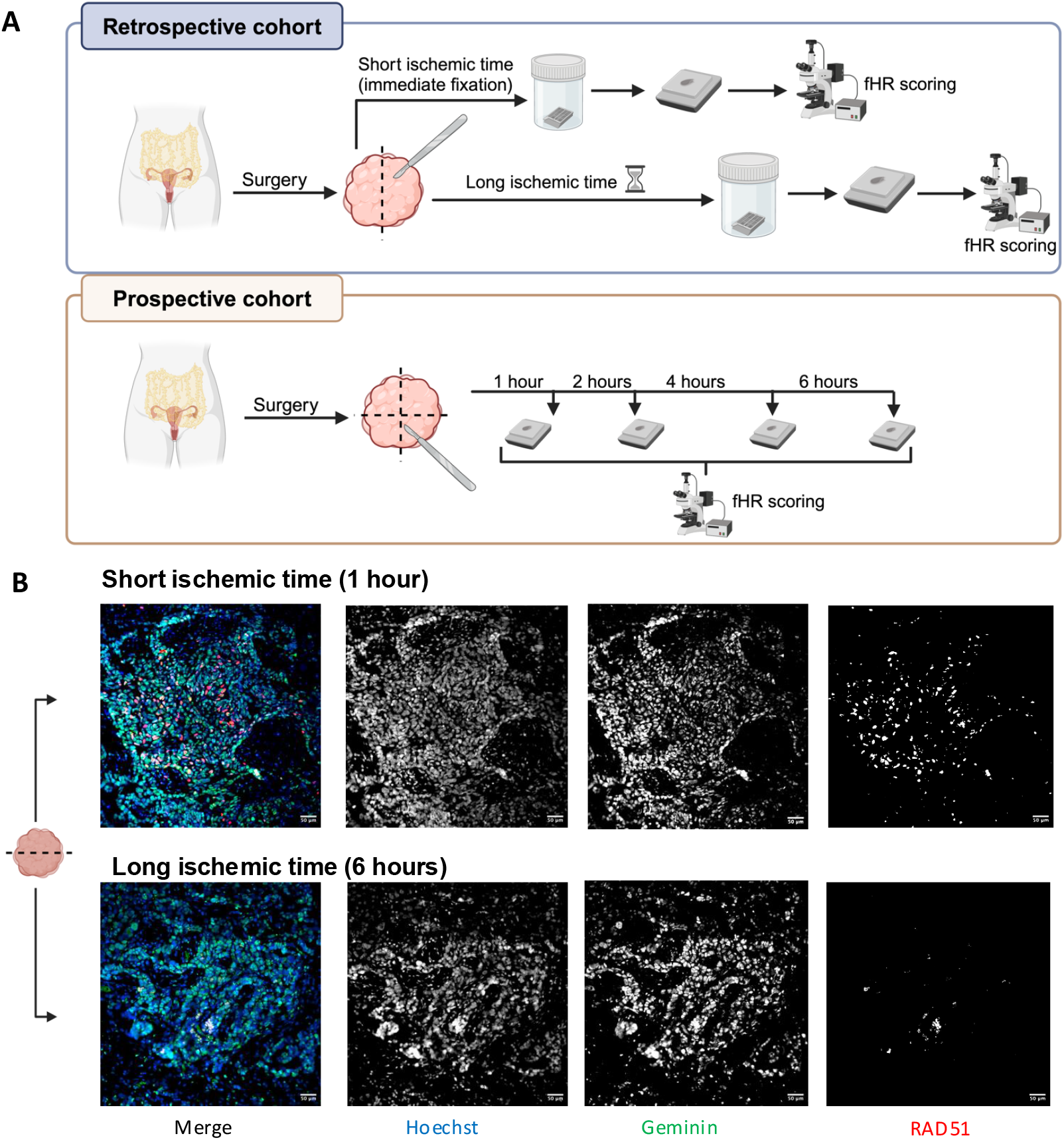
Study design and experimental strategy. **A**. Sample processing workflow. Each tumor sample was divided into two or more pieces, one of which was delivered into fixative quickly and the other(s) with a delay. After paraffin embedding, samples were subjected to functional HR testing and categorized as functionally HR-deficient (fHRD) or HR-proficient (fHRP) (Pikkusaari 2023). **B**. Representative images of immunofluorescence staining for fHR scoring, after long and short ischemic times. Samples are from the same tumor specimen, with fixation performed after ischemic times of different lengths. The tumor sample demonstrates a significant reduction in the RAD51 signal (a marker for HR-mediated DNA repair) after a long ischemic period of 6 hours. Scale bars =50μm.

#### Retrospective cohort

The retrospective cohort consisted of 18 retrospectively analyzed chemo-naive samples from 15 patients in the DECIDER study, who underwent primary debulking surgery or diagnostic laparoscopy followed by neo-adjuvant chemotherapy treatment at Turku University Central Hospital (Supplementary Table 1). Pre-existing archival paired FFPE blocks originating from the same tumor were used, one with short and one with long ischemic time. The former tumor piece was fixed on site in 10% formalin immediately after surgical excision (ischemic time of <2 hours), while the latter underwent transportation from Turku University Hospital to our lab at the University of Helsinki in saline buffer, before being fixed in 10% formalin, resulting in an ischemic time of approximately four hours (Figure 1A).

#### Prospective cohort

The prospective cohort included 9 PDS samples from 9 patients in the ONCOSYS-OVA study at Helsinki University Hospital (Supplementary Table 1). To determine the maximum ischemic time for reliable and accurate RAD51 protein quantification, each HGSC tumor sample was divided into four segments and left at room temperature in saline buffer for 1, 2, 4, or 6 hours before they were fixed in 10% formalin (Figure 1A). This time course aimed to mimic the range of ischemic times encountered in current HGSC clinical diagnostics.

## Immunofluorescence staining

FFPE samples were sectioned, deparaffinized in xylene, and rehydrated through an ethanol series. Sections were boiled in citrate buffer (pH 6) for 15 minutes to retrieve antigens. Afterwards, the sections were stained with primary antibodies against γH2AX (Abcam Cat# ab11174, dilution 1:1000), geminin (Abcam Cat# ab104306, dilution 1:500), and RAD51 (Abcam Cat# ab133534, dilution 1:1000). For each sample, two adjacent sections were used, each stained with one of the following combinations: geminin+γH2AX or geminin+RAD51. The sections were then treated with fluorescently labeled secondary antibodies; donkey anti-mouse IgG-AlexaFluor™ 488 (Invitrogen Cat# A-21202, 1:500 dilution) and donkey anti-rabbit IgG-AlexaFluor™ 647 (Invitrogen Cat# A-31573, 1:500 dilution) and counterstained with Hoechst to visualize nuclei. Finally, the slides were scanned at 20x magnification using a Pannoramic 250 FLASH III slide scanner.

### Functional HR scoring

Functional HR (fHR) capacity was assessed as described previously (Pikkusaari 2023). Briefly, regions of interest from images of IF-stained FFPE sections were selected based on γH2AX and geminin staining, to ensure that regions with sufficient levels of S-phase DNA damage were analyzed. Analysis was conducted semi-automatically in ImageJ with custom macros. Tumor cell nuclei were identified by their size using Hoechst staining. Mask of tumor nuclei was applied to the geminin channel to identify S/G2 phase nuclei. DNA damage and HR-mediated repair were quantified by assessing γH2AX and RAD51, respectively, in S/G2 phase nuclei, with particle counting used for RAD51 and γH2AX-positive nuclei. fHR scores were calculated as a percentage of RAD51-positive nuclei out of all S/G2 phase nuclei. Samples were categorized as either fHRD or functionally HRP (fHRP) based on the cutoff value of 10%, as determined previously on HGSC samples (those with a value of 10% or below as fHRD, and >10% as fHRP; Pikkusaari 2023).

### tatistical Analyses

Statistical analyses for all data were performed using GraphPad Prism 9 software (GraphPad Software, San Diego, CA, USA, www.graphpad.com). The statistical test used for each analysis is provided in the figure legends, with a P value of < 0.05 considered statistically significant.

## Results

In this study, we assessed the impact of variable ischemic time on fHR scores in a total of 27 HGSC samples. Using nuclear RAD51 positivity as a marker, we quantified the fHR scores to evaluate how ischemic length impacts biomarker detection in these tumor samples. In the ischemic time course performed on the prospective cohort, cells with nuclear RAD51 signal drastically decreased after 6 hours of ischemic time compared to 1 hour of ischemic time of the same tumor (Figure 1B). Longer ischemic times had no impact on IF staining for geminin or γH2AX (Supplementary Figure 1). In the retrospective cohort, 12 out of 18 samples had lower fHR scores after prolonged ischemic time, while 5 samples exhibited no significant change (Figure 2A). Notably, upon a long ischemic time, 6/18 samples (33 %) of the retrospective-cohort transitioned from the fHRP category to the fHRD category, i.e. their fHR score dropped below the 10% cut-off value.

**Figure 2.**
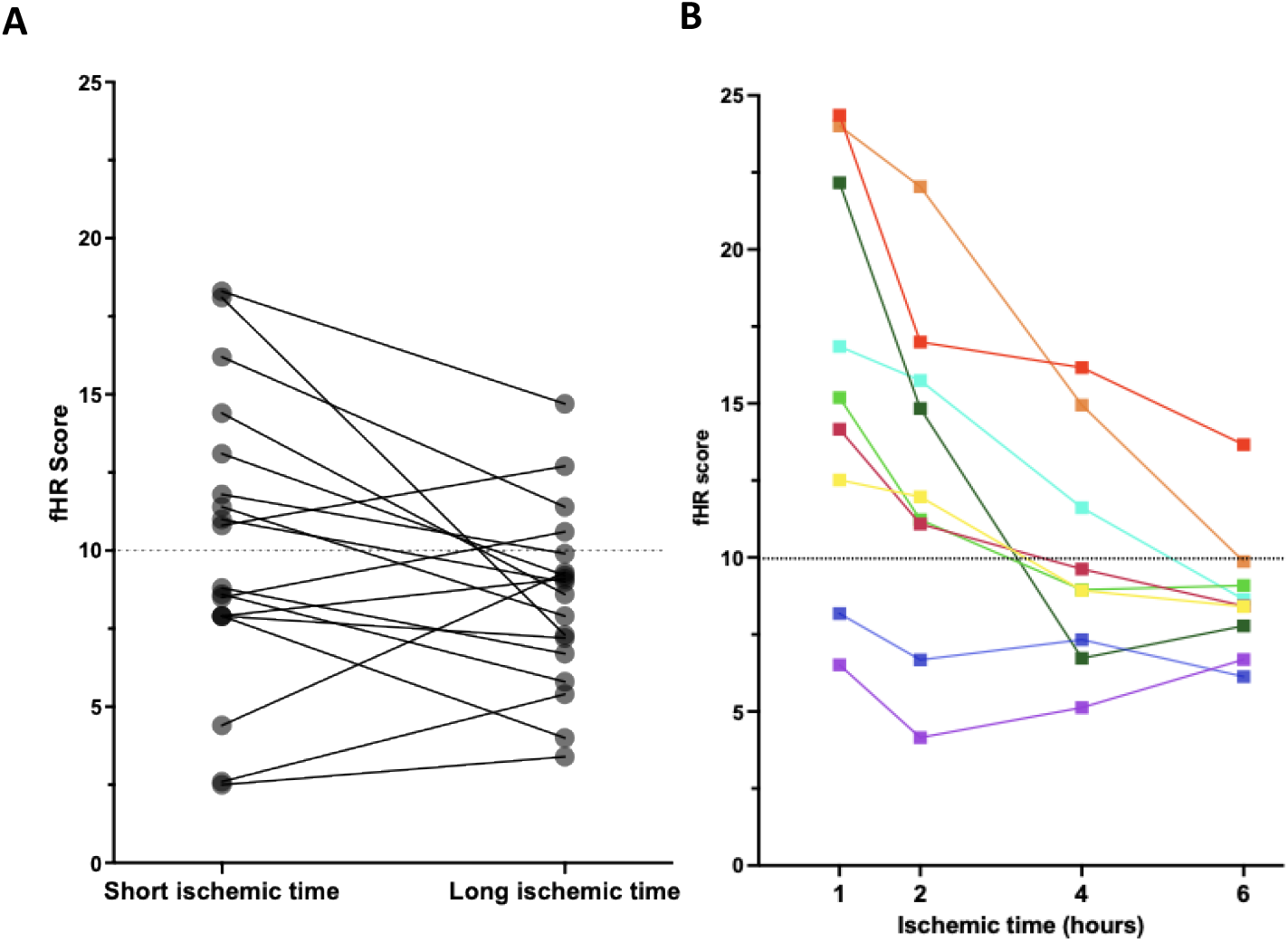
Longer ischemic times result in decreased fHR scores, i.e. potentially in false fHRD calls. Dashed lines indicate the cut-off value for fHRD (below 10 is fHRD in chemo-naïve samples, Pikkusaari 2023). **A)** The impact of delayed fixation on fHR scores was retrospectively determined in 15 HGSC patients for whom two pieces of the same tumor, one with short (1 hour) and one with long (4 hours) ischemic time, were available (n=18 samples). **B)** Systematic testing of the impact of ischemic time on fHR score in an independent, prospective cohort of 9 HGSC patients. Each HGSC tumor (n=9 samples) was cut into four pieces that were then delivered into fixative at four different time points (1–6 hours after surgical resection).

In the prospective cohort, fHR scores decreased in all 9 samples already after 2 hours of ischemic time compared to 1 hour (Figure 2B). For two samples initially (at the 1-h time point) categorized as fHRD, there was no overall decrease in fHR score with longer ischemic time, though slight changes were observed (Sample #1: median fHR score 5.8, range 4.1–6.7; and Sample #2: median fHR score 7.0, range 6.1–8.2). The ischemic time of 4 hours led to a change in the fHR category in 4/9 samples, changing the fHR classification from fHRP to fHRD. After a prolonged ischemic time of 6 hours at room temperature, 5/9 samples in total had shifted from fHRP to fHRD.

We then combined data from the two cohorts for a more comprehensive assessment of the impact of ischemic time. To enable comparison and to align the two cohorts, we classified samples in the prospective cohort with a 1-hour ischemic time as “short” and those with a 4-hour ischemic time as “long”. fHR scores were significantly lower in the long ischemic time group than in the short ischemic time group. The fHR scores derived from samples with short ischemic times demonstrated greater variability (range: 2.5–23; median: 11.4) compared to those derived from samples with long ischemic times (range: 4–14.9; median: 9) (Figure 3). Notably, the distribution of fHR scores from samples with short ischemic times were more similar to the findings from our previous study, which reported a range of 1.8–23.8 in the validation cohort (Pikkusaari, 2023).

**Figure 3.**
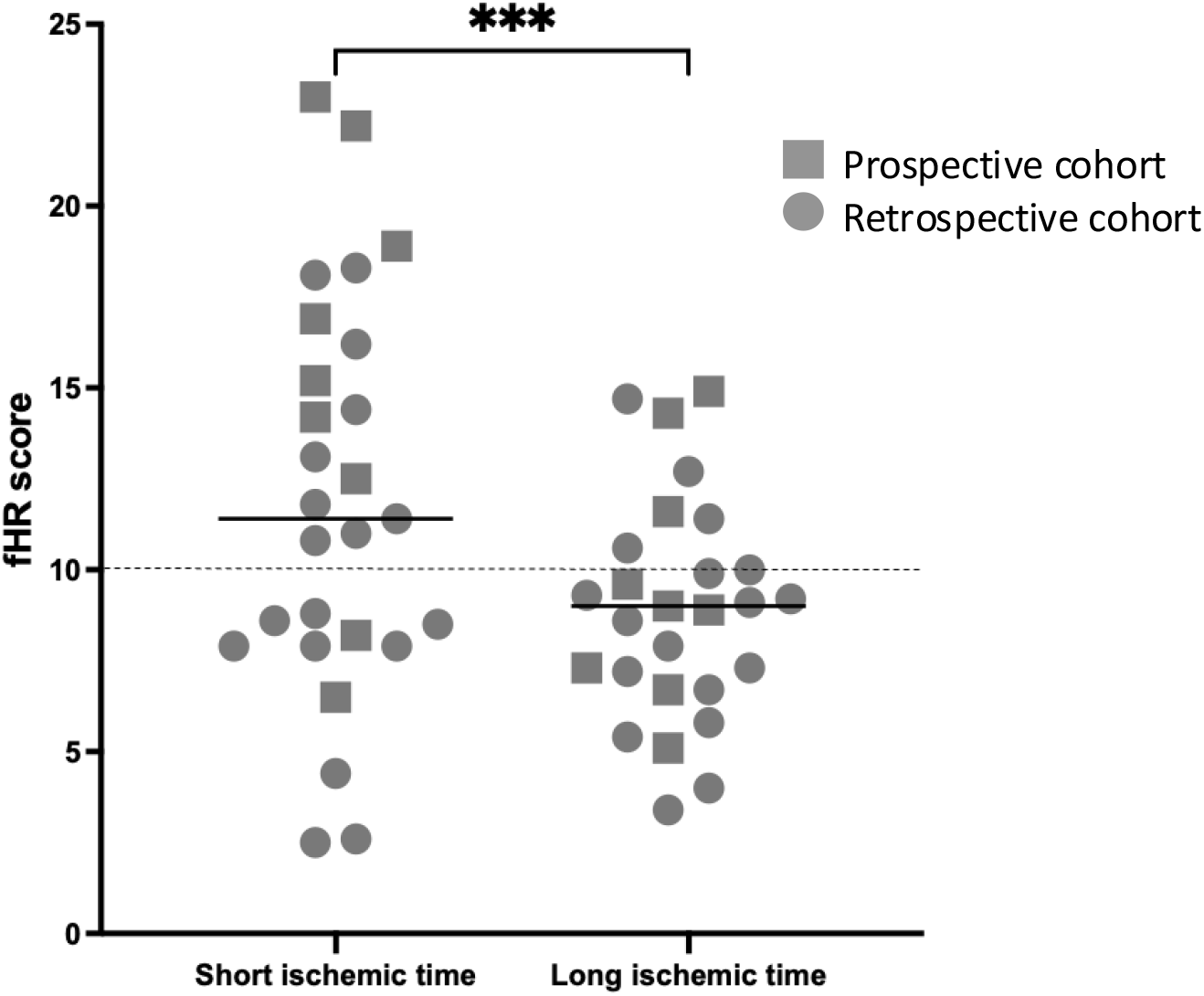
Functional HR (fHR) scores of 27 samples (retrospective and prospective cohort combined), each with short and long ischemic times. Black horizontal lines indicate mean fHR values for each group of samples. The dashed line marks the cut-off value for fHRD (below 10 is fHRD in chemo-naïve samples, Pikkusaari et al. 2023). fHR scores were significantly different between the two groups (***p<0.001, Wilcoxon test)

Further, we asked whether prolonged ischemic time (4 hours versus 1 hour) differentially impacted fHR scores derived from primary vs. metastatic tumor sites (Figure 4). In samples from primary tissue sites, the fHR status of 7/12 samples did not change even after a long ischemic time. In contrast, 11 out of 22 samples from metastatic sites changed their HR status following extended ischemic time. Metastatic (omental and peritoneal tumor) samples thus appear more prone to a substantial drop in fHR scores upon extended ischemic time.

**Figure 4.**
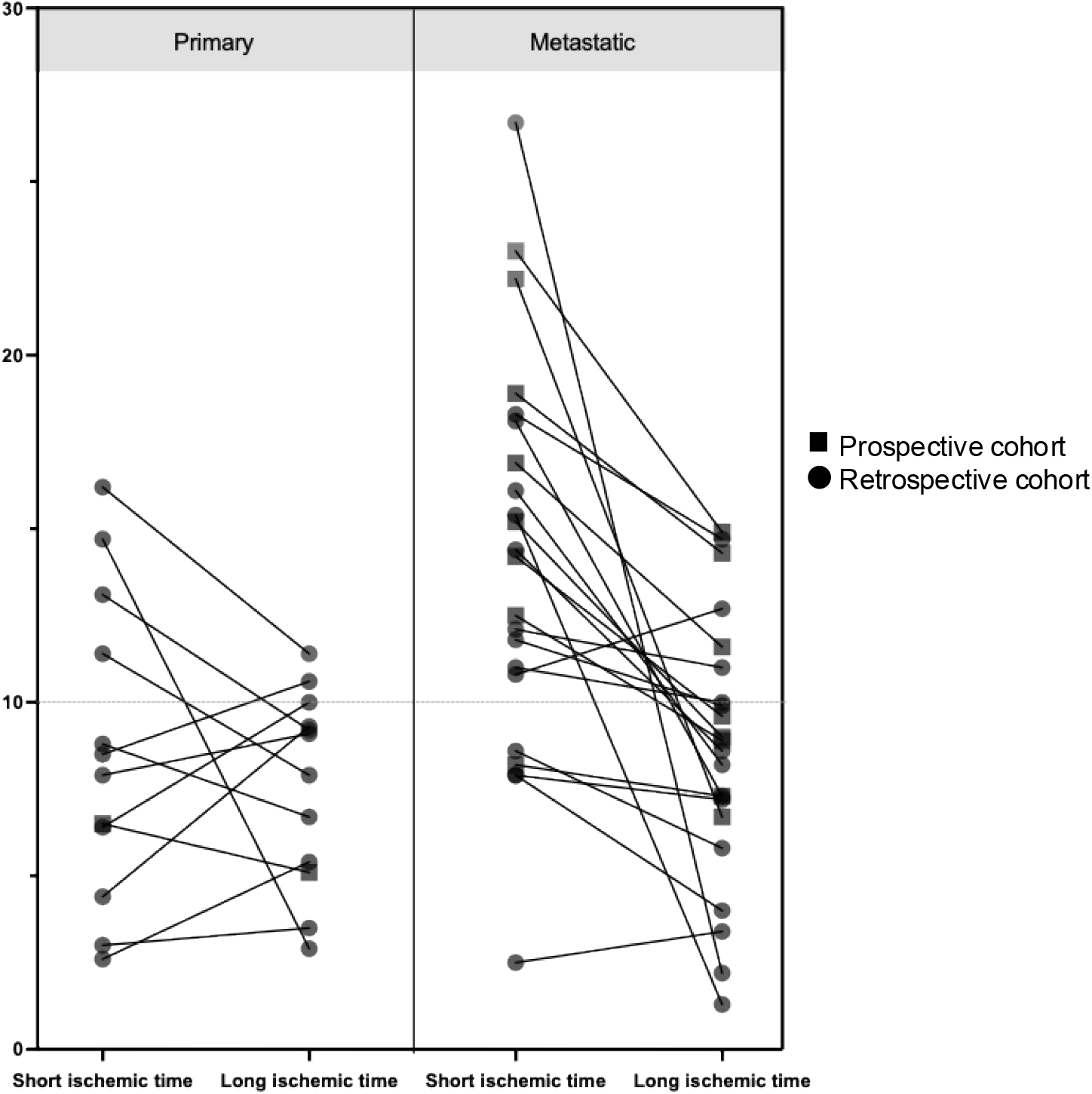
Prolonged ischemic time had a greater impact on fHR score in metastatic tumor specimens compared to primary tumor specimens. Primary tissue sites include: ovary, and fallopian tubes. Metastatic tissue sites include omentum and peritoneum. Dashed horizontal lines mark the cut-off value for fHRD (below 10 is fHRD in chemo-naïve samples, Pikkusaari et al. 2023). For primary tissue site samples, fHR scores were more similar between the time points. The score in most of the samples obtained from metastatic tissue sites, fHR scores decreases substantially after long ischemic time.

As part of fHR scoring, the percentage of geminin-positive (S/G2 phase) cells was calculated. Unlike the RAD51 signal, we found no substantial, consistent decrease in the geminin signal with prolonged ischemic time (Supplementary Fig. 1A) (see also example immunofluorescence images in Figure 1B). In addition to RAD51 and geminin, we also included an analysis of γH2AX positivity, since quantifying nuclear γH2AX is a pre-requisite for fHR scoring. No evident difference in the amount of total DNA damage (% γH2AX-positive cells) was found between different ischemic times within the same tumor sample (Supplementary Fig. 1B).

## Discussion

Reliable HRD calling is essential to tailoring effective treatment strategies for HGSC patients. Functional biomarkers of HR capacity, such as RAD51, have great untapped potential to guide PARP inhibitor treatment decisions (Miller 2020). In this study, we investigated the impact of ischemic time on DNA repair marker RAD51 in HGSC specimens, both empirically and experimentally. Our findings underscore ischemic time as a key pre-analytical variable that can influence the integrity of HGSC biomarkers. Numerous studies have examined the impact of pre-analytical variables on immunohistochemical markers, such as time spent in fixative, type of fixative (including concentration, pH, etc.), choice of antibody and concentration, and slide-drying as well as analytical variables, including scoring methods and interobserver variability (Khoury 2018, Goldstein 2003, Cohen 2012, Potts 2012). However, the impact of these factors in protein detection has not been fully investigated in HGSC.

Our results demonstrate that delayed time-to-fixation can significantly alter the ability to detect DNA repair marker RAD51 in HGSC FFPE specimens, leading to false-positive HRD calls. Based on the RAD51 quantification in samples that experienced short and long ischemic times, a tolerable upper limit for ischemic time for fHR scoring appears to be 2 hours. This is consistent with ASCO-CAP guidelines, which recommend keeping ischemic time under 2 hours when collecting FFPE samples for IHC assays, to ensure the validity of biomarker assessment (Hammond 2010).

In their evaluation of RAD51-FPPE test performance, van Wijk et al. (2023) used previously defined parameters to examine the RAD51 biomarker in breast cancer tissue specimens. Despite efforts to limit ischemic time to two hours, the authors acknowledged potential variances in tissue quality due to the diagnostic nature of specimen collection. Specifically, they also hypothesized that longer ischemic times in some samples might contribute to false-positive HRD results (van Wijk 2023).

Our findings also align with literature that indicates the sensitivity of breast cancer biomarkers to ischemic degradation, where 30-minute cold ischemic times affected immunohistochemical staining for progesterone receptors and a 2-hour delay impacted both hormone receptors and HER2 expression (Yildiz-Aktas et al. 2012). Similarly, in liver tumors, Lerch et al. (2020) observed improvements in the preservation of phosphoproteins when reducing ischemic time and using cold collection transport devices. Further, degradation of the tissue samples upon long ischemic time was shown to result in a significant reduction of expression levels of biomarkers like Ki-67, a commonly used marker of tumor proliferation ability (Arima 2015).

We also used the γH2AX signal (which is required in fHR testing for calculating the DNA damage) as a comparison to the RAD51 signal, since both are nuclear markers. Except for a few outliers, there was no notable difference in total DNA damage. γH2AX is the phosphorylated form of the histone protein H2AX, a structural component of chromatin (Kuo 2008). As a result, γH2AX is more compact and structurally stable. One hypothesis is that this localization within the chromatin provides greater protection, making yH2AX less affected by ischemic time compared to RAD51. The variation in the scores in different ischemic times may stem from technical inconsistencies or tumor heterogeneity within a sample (Supplementary Fig. 1B). This could also explain the slight variation in the percentage of geminin-positive cells.

Functional HRD testing is cost-effective and has substantial utility for clinical diagnostics in the future, especially for samples with low tumor cell content where genomics-based assays fail. Moreover, because it is agnostic to the underlying cause of tumor HR deficiency (mutational or otherwise), it not only circumvents the problem of interpreting variants of unknown significance (VUS) in HR genes, but in fact can be used to test whether the identified VUSs lead to inactivation of HR in the tumor. Functional HRD testing can also identify cases with BRCA reversions that restore HR capacity (Pikkusaari 2023). Thus, fHRD testing would be a valuable addition to the clinical HRD diagnostics toolbox. To ensure accurate fHRD calls in HGSC tumors, ischemic time as a key preanalytical variable needs to be controlled for. In the hospital pathology units, long ischemic times (even >4 hours) are not uncommon especially for HGSC cases where surgery is long and complex. Our findings highlight the need for robust, standardized protocols for HGSC tissue handling in clinical and research settings. Recording exact tissue collection times and the initiation of fixation would mitigate variability in biomarker assessments. For archival tumor samples (e.g. biobank specimens) where such information is often lacking, caution is warranted when interpreting immunostaining results obtained from them, be it for clinical biomarker discovery or for cancer research purposes. While this study did not measure RNA-level readouts in samples with extended ischemic times, it is reasonable to assume that FFPE-based transcriptomics would be impacted even more severely, given that RNA molecules a much more labile than proteins. Variability in pre-analytical steps, if not accounted for, could profoundly affect spatial biology studies of cancer.

In conclusion, this study highlights the critical need to consider ischemic time as part of tumor sampling protocols, particularly for FFPE samples used in protein biomarker analyses like IHC assays. Rapid and standardized fixation protocols should be established, ideally delivering tissue samples into fixative within 2 hours post-resection to minimize ischemic degradation.

## Data Availability

All data produced in the present study are available upon reasonable request to the authors

## Acknowledgements

We wish to thank study nurses Laura Haapasalo and Maija Vääriskoski. We also thank Minna Eriksson, Sweta Jha and Anastasia Lundgren for their excellent technical assistance.

We thank the tissue preparation and histochemistry unit of the Anatomy Department at University of Helsinki. We thank the Digital Microscopy and Molecular Pathology unit at the Institute of Molecular Medicine Finland (FIMM) for the histology services provided and the whole-slide scanning services provided by the Genome Biology Unit, HiLIFE, University of Helsinki.

This study was funded by the Cancer Foundation Finland (L.K.), the Sigrid Juselius Foundation (L. K.), the European Union’s Horizon 2020 program to (grant number 965193, DECIDER project) and the Research Council of Finland’s iCAN Digital Precision Cancer Medicine Flagship project.

