## Supplementary Data for "Length of ischemic time is critical for accurate determination of homologous recombination capacity by immunostaining in FFPE tumor samples"

| | | retrospective cohort | prospective cohort |
|---|---|---|---|
| Total cases | n | 15 | 9 |
| Median age at diagnosis (range) | years | 68<br>(51-83) | 63,5<br>(48-76) |
| FIGO stage | III | 10 | 2 |
| | IV | 5 | 7 |
| Treatment strategy | PDS | 13 | 8 |
| | NACT | 2 | 1 |
| Primary therapy response | CR | 9 | 5 |
| | PR | 6 | 1 |
| | PD | - | 1 |
| | unknown | - | 2 |
| Cytoreduction | R0 | 8 | 1 |
| | R>0 | 7 | 8 |
| PARPi | frontline | 5 | 1 |
| | recurrence | 3 | - |
| | none | 7 | 8 |

**Supplementary Table 1.** Clinicopathological characteristics of patients included in the study, PDS: primary debulking surgery, NACT: neo-adjuvant chemotherapy, CR: Complete response, PR: Partial response, PD: Progressive disease

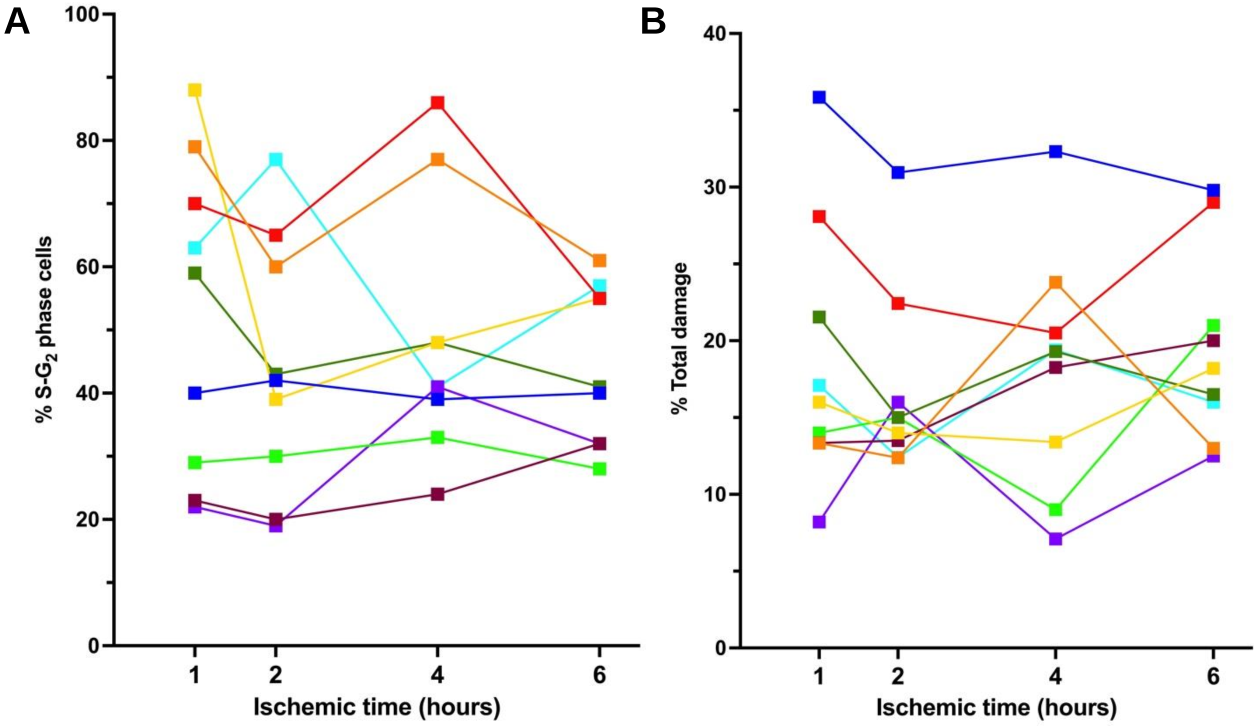


**Supplementary figure 1. Quantification of positivity for additional markers after prolonged ischemic time.**

**A)** Geminin-positive (S-$G_2$ phase) nuclei out of total nuclei, at various ischemic time points. There was no substantial or consistent decrease in geminin signal. **B)** γH2AX-positive nuclei (DNA damage) out of total nuclei, at various ischemic time points.
